# Degree of Cyclooxygenase-2 Inhibition Modulates Blood Pressure Response to Celecoxib and Naproxen

**DOI:** 10.1101/2024.05.30.24308244

**Authors:** Katherine N. Theken, Soumita Ghosh, Carsten Skarke, Susanne Fries, Nicholas F. Lahens, Dimitra Sarantopoulou, Gregory R. Grant, Garret A. FitzGerald, Tilo Grosser

## Abstract

**Background:** Non-steroidal anti-inflammatory drugs (NSAIDs) increase the risk of adverse cardiovascular events via suppression of cyclooxygenase (COX)-2-derived prostacyclin (PGI_2_) formation in heart, vasculature, and kidney. The Prospective Randomized Evaluation of Celecoxib Integrated Safety versus Ibuprofen Or Naproxen (PRECISION) trial and other large clinical studies compared the cardiovascular risk of traditional NSAIDs (i.e. naproxen), which inhibit both COX isozymes, with NSAIDs selective for COX-2 (i.e. celecoxib). However, whether pharmacologically equipotent doses were used - that is, whether a similar degree of COX-2 inhibition was achieved - was not considered. We compared drug target inhibition and blood pressure response to celecoxib at the dose used by most patients in PRECISION with the lowest recommended naproxen dose for osteoarthritis, which is lower than the dose used in PRECISION.

**Methods:** Sixteen healthy participants (19-61 years) were treated with celecoxib (100 mg every 12h), naproxen (250 mg every 12h), or placebo administered twice daily for seven days in a double-blind, crossover design randomized by order. On Day 7 when drug levels had reached steady state, the degree of COX inhibition was assessed *ex vivo* and *in vivo*. Ambulatory blood pressure was measured throughout the final 12h dosing interval.

**Results:** Both NSAIDs inhibited COX-2 activity relative to placebo, but naproxen inhibited COX-2 activity to a greater degree (62.9±21.7%) than celecoxib (35.7±25.2%; p<0.05). Similarly, naproxen treatment inhibited PGI_2_ formation *in vivo* (48.0±24.9%) to a greater degree than celecoxib (26.7±24.6%; p<0.05). Naproxen significantly increased blood pressure compared to celecoxib (differences in least-square means of mean arterial pressure: 2.5 mm Hg (95% CI: 1.5, 3.5); systolic blood pressure: 4.0 mm Hg (95% CI: 2.9, 5.1); diastolic blood pressure: 1.8 mm Hg (95% CI: 0.8, 2.8); p<0.05 for all). The difference in systolic blood pressure relative to placebo was associated with the degree of COX-2 inhibition (p<0.05).

**Conclusions:** Celecoxib 200 mg/day inhibited COX-2 activity to a lesser degree than naproxen 500 mg/day, resulting in a less pronounced blood pressure increase. While the PRECISION trial concluded the non-inferiority of celecoxib regarding cardiovascular risk, this is based on a comparison of doses that are not equipotent.

ClinicalTrials.gov identifier: NCT02502006 (https://clinicaltrials.gov/study/NCT02502006)

**Clinical Perspective:**

- Naproxen 250 mg twice a day inhibited COX-2 activity to a greater degree than celecoxib 100 mg twice a day.
- The degree of COX-2 inhibition was associated with the increase in systolic blood pressure with NSAID treatment relative to placebo.
- Dose and its pharmacological potency achieved *in vivo* should be considered when evaluating the relative cardiovascular safety of COX-2-selective vs. non-selective NSAIDs.

## Introduction

Non-steroidal anti-inflammatory drugs (NSAIDs) are among the most commonly used medications worldwide and the opioid crisis has placed a new emphasis on their use.^1,2^ Approximately 20% of adults in the United States receive at least one NSAID prescription per year^3^, and 12% of adults in the United States reported using NSAIDs chronically, i.e. more than three times weekly for more than three months.^4^ Consumption of NSAIDs in individuals at risk for musculoskeletal injuries is even more common.^5-7^ Given the high prevalence of chronic pain in the United States – more than 100 million Americans suffer from chronic pain^8^ – NSAIDs are an important non-addictive option for pain relief. Optimizing NSAID therapy is one strategy to address the current opioid crisis,^2^ and rates of NSAID use have increased in recent years as opioid prescriptions have declined.^9^ Although they lack the addictive potential of opioids, NSAIDs have the potential to cause serious, and in some cases, life-threatening adverse events, including gastrointestinal bleeding, renal dysfunction, hypertension, and thrombotic cardiovascular events.^10^ Undetected blood pressure (BP) increases that may impact cardiovascular morbidity and mortality at the population level are a particular concern.^11^

NSAIDs exert their analgesic and anti-inflammatory effects via inhibition of cyclooxygenase (COX)-1 and/or COX-2, enzymes that catalyze the first committed step in prostaglandin (PG) synthesis. PGs produce a diverse array of biologic effects via activation of prostanoid receptors and play important roles in a variety of pathologic and homeostatic processes.^12^ The risk of thrombotic events associated with the use of NSAIDs, particularly those selective for COX-2, is mediated via suppression of COX-2-derived prostacyclin (PGI_2_) formation in endothelial and vascular smooth muscle cells.^13,14^ PGI_2_ possesses potent anti-thrombotic and vasodilatory effects, and thus acts as a general inhibitor of platelet activation *in vivo*.^12^ Non-selective NSAIDs also inhibit COX-2 in the vasculature, but the associated risk of thrombosis is mitigated to some extent by inhibition of formation of thromboxane A_2_ (TxA_2_), a COX-1-derived PG released by activated platelets that promotes platelet activation and aggregation.^10,15^ In addition to their effects on vascular PG production, NSAIDs inhibit renal PG formation, resulting in sodium retention and BP increases, which may further augment cardiovascular risk.^10,15,16^

One of the largest (N=24,081) outcome studies to date, the Prospective Randomized Evaluation of Celecoxib Integrated Safety versus Ibuprofen Or Naproxen trial (PRECISION), compared the safety of the COX-2 selective NSAID, celecoxib (100 to 200 mg twice a day), and two traditional NSAIDs, naproxen (375 to 500 mg twice a day) and ibuprofen (600 to 800 mg three times a day), in osteoarthritis (90%) and rheumatoid arthritis (10%) patients. PRECISION concluded that the cardiovascular safety of moderate doses of celecoxib (average dose: 209±37 mg/day) was noninferior to naproxen (average dose: 852±103 mg/day) or ibuprofen (average dose: 2045±246 mg/day).^17^ However, a secondary on-treatment analysis of PRECISION showed a lower risk of cardiorenal events in the celecoxib group than the ibuprofen and naproxen groups.^18^ Similarly, a pre-specified substudy (PRECISION-ABPM) reported that the percentage of patients with normal baseline blood pressure who developed hypertension was significantly greater in patients treated with naproxen (19%) or ibuprofen (23%), than with celecoxib (10%).^19^ However, the degree of COX-2 inhibition attained has never been assessed in clinical outcome trials, and thus it is unknown whether the doses employed in PRECISION were equipotent. Here, we compared the pharmacologic potency of celecoxib, 200 mg/day (the highest dose allowed for osteoarthritis and roughly the average daily dose in PRECISION), with naproxen, 500 mg/day (the lowest recommended dose for osteoarthritis and approximately 40% lower than the average daily dose in PRECISION), and how this relates to blood pressure response to NSAIDs in a highly controlled study in apparently healthy volunteers.

## Methods

### Participants

Men and women (≥18 years of age) who were in good health based on medical history, physical examination, vital signs, and laboratory tests were enrolled. Participants were excluded if they were pregnant or nursing a child, smoked or used nicotine-containing products, were obese (body mass index (BMI) > 30 kg/m^2^), had a history of significant cardiovascular, gastrointestinal, renal, hepatic, respiratory, immune, endocrine, hematologic, or neurological disease, a history of cancer within the last 5 years, or a coagulation or bleeding disorder, were sensitive or allergic to celecoxib, naproxen, aspirin, or other NSAIDs, or had used NSAIDs (including acetaminophen), dietary or herbal supplements containing salicylates, Vitamin E, fish oil, or any other herbal supplements, within 14 days of study drug administration.

### Study procedures

The study protocol was approved by the University of Pennsylvania Institutional Review Board (IRB#820715; ClinicalTrials.gov: NCT02502006), and all participants provided informed consent. The study was a randomized, double-blind, three-way crossover study comparing the degree of COX inhibition and the blood pressure response at steady state following treatment with celecoxib (100 mg twice daily), naproxen (250 mg twice daily), or placebo (twice daily) for 7 days. Prior to beginning treatment, all participants attended a screening visit to obtain a complete medical history and confirm eligibility. They were asked to abstain from analgesics, including products containing NSAIDs (including aspirin and acetaminophen), high dose vitamins, and nutritional supplements until study completion.

On the first day of each treatment phase, baseline blood and urine samples were collected, and participants were given a blisterpack with blinded study medication to be taken by mouth twice daily on an outpatient basis. Study medication was blinded by over-encapsulation by the University of Pennsylvania Investigational Drug Service. On the morning of day 7, participants returned to clinic for a 12-hour visit for pharmacokinetic-pharmacodynamic (PK-PD) sampling. An ambulatory blood pressure monitor (ABPM, Spacelabs 90207), which recorded blood pressure every 15 minutes throughout the study visit, was placed on the non-dominant upper arm. Blood and urine samples were collected (T=0), and the final dose of study medication was administered. Additional samples were collected 0.5 (blood only), 1, 2, 4, 8, and 12 hours after study medication administration. Participants were discharged after the 12-hour sample collection. These study visits were repeated for the next two treatment phases, with a washout period of at least 2 weeks between each treatment phase.

Study data were collected and managed using REDCap (Research Electronic Data Capture) hosted at the University of Pennsylvania Perelman School of Medicine.^20,21^

### Quantification of COX activity and plasma drug concentrations

COX-1 activity *ex vivo* was evaluated by quantifying serum thromboxane B_2_ levels, as previously described.^22^ Briefly, whole blood was collected into vacuum tubes containing clot activator and incubated in a water bath at 37°C for 1 hour. Serum was separated by centrifugation and stored at -80°C until analysis by liquid chromatography-tandem mass spectrometry (LC-MS/MS).

COX-2 activity *ex vivo* was evaluated by quantifying plasma PGE_2_ levels following lipopolysaccharide (LPS) stimulation in whole blood, as previously described.^23^ Briefly, heparinized whole blood was treated with aspirin (1 mM) and incubated at room temperature for 15 minutes. LPS (E. coli, serotype O111:B4, 10 µg/ml whole blood) was added, and the sample was incubated in a water bath at 37°C for 24 hours. Plasma was separated by centrifugation and stored at -80°C until analysis by LC-MS/MS.

COX activity *in vivo* was determined by quantification of urinary prostanoid metabolites by LC-MS/MS as previously described.^14^ Systemic production of PGI_2_, PGE_2_, PGD_2_, and thromboxane (Tx) A_2_ was determined by quantifying their major urinary metabolites: 2,3-dinor 6-keto-PGF_1α_ (PGIM), 7-hydroxy-5,11-diketotetranorprostane-1,16-dioic acid (PGEM), 11,15-dioxo-9α-hydroxy-2,3,4,5-tetranorprostan-1,20-dioic acid (PGDM), and 2,3-dinor TxB_2_ (TxM), respectively. Results were normalized to urinary creatinine measured by LC-MS/MS.

Plasma concentrations of celecoxib and naproxen were quantified by LC-MS/MS as previously described.^24^

### Statistical analysis

Measurements of COX activity *ex vivo* and urinary PG metabolite levels were normalized to the mean value during the placebo phase for each subject to calculate the percent of COX inhibition relative to placebo. Area-under-the-curve from T=0 to T=12 hours (AUC) was calculated as a measure of the degree of COX inhibition throughout the dosing interval. The degree of COX inhibition was compared by paired t-test. The effect of treatment on COX activity and ABP over time was analyzed by linear mixed effect modeling using the lme4 R package,^25^ including time and treatment as main effects and participant as a random effect. The relationship between change in SBP and COX inhibition over the 12-hour dosing interval was evaluated by linear regression. P<0.05 was considered statistically significant. Statistical analyses were performed in R (version 4.3.1).

## Results

The study cohort included 16 healthy adults (9 men, 7 women) with a mean age of 34.7±13.4 years. Baseline demographic and clinical characteristics are shown in Table 1. Biochemical measures were evaluated in all participants. One participant was excluded from ABP analysis due to ABPM malfunction and incomplete data.

**Table 1.** Baseline characteristics of study participants.

| Characteristic | Study Cohort |
| --- | --- |
| N (Men/Women) | 16 (9/7) |
| Age | 34.7±13.4 years (range: 19-61 years) |
| Race |  |
| White | 9 |
| Black/African American | 4 |
| Asian | 1 |
| Other | 2 |
| Ethnicity |  |
| Non-Hispanic | 13 |
| Hispanic | 3 |
| Body mass index | 22.6±1.9 kg/m<sup>2</sup> |
| Lab values |  |
| Total cholesterol | 166.9±27.6 mg/dl |
| Triglycerides | 70.9±24.4 mg/dl |
| LDL | 94.7±22.9 mg/dl |
| HDL | 58.1±13.3 mg/dl |
| Serum creatinine | 0.83±0.14 mg/dl |
| Systolic blood pressure | 115.3±8.9 mm Hg |
| Diastolic blood pressure | 69.8±9.5 mm Hg |
| Heart rate | 65.7±9.8 bpm |

Naproxen 250 mg twice daily inhibited COX-1 activity and COX-2 activity *ex vivo* by 95.3±4.4% and 62.9±21.7%, respectively, while celecoxib 100 mg twice daily had minimal effects on COX-1 activity *ex vivo* and inhibited COX-2 activity *ex vivo* by 35.7±25.2% over the 12-hour dosing interval (Figure 1). Similar results were observed for COX activity *in vivo*, assessed by urinary PG metabolite concentrations. TxM (an index of COX-1 activity) was inhibited by 68.2±18.7% with naproxen treatment and 8.9±35.7% with celecoxib treatment. PGIM (an index of COX-2 activity *in vivo*) was inhibited by 48.0±24.9% with naproxen treatment and 26.7±24.6% with celecoxib treatment. With all functional parameters, the degree of COX inhibition was significantly greater with naproxen treatment than celecoxib treatment (p<0.05). The maximum plasma concentration (C_max_) of naproxen was 228±45.1 µM, and the time to maximum plasma concentration (t_max_) was 1.8±1.2 h after administration. For celecoxib, C_max_ was 1.28±0.55 µM, and t_max_ was 2.3±1.2 h.

**Figure 1.**
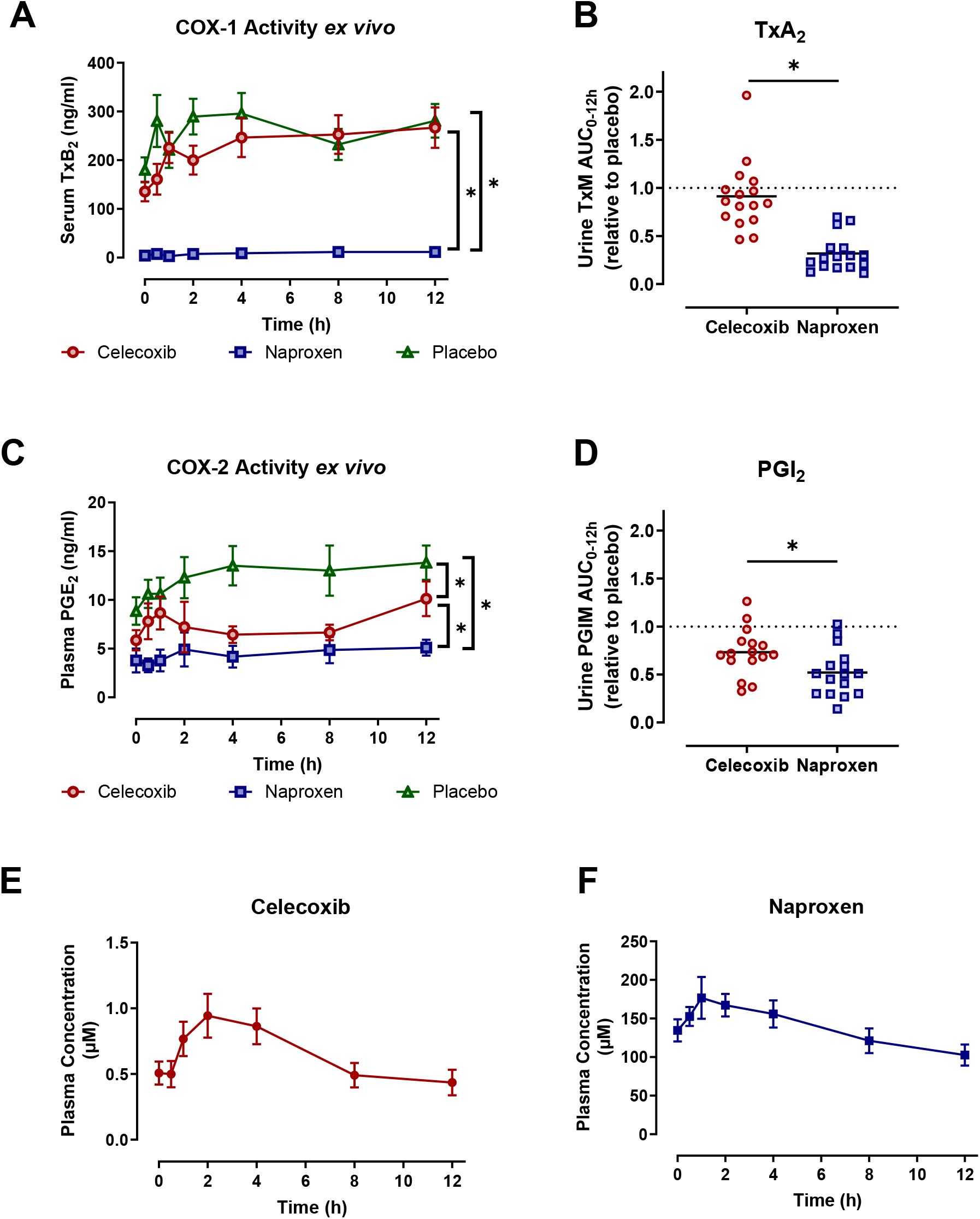
Comparison of (A) COX-1 inhibition *ex vivo* and (B) *in vivo* and (C) COX-2 inhibition *ex vivo* and (D) *in vivo* by treatment. Plasma concentrations of (E) celecoxib and (F) naproxen over time. Dosing occurred at 8:00 AM ± 15 min. Data are shown as mean ± SEM. *p<0.05

During the placebo phase, average MAP, SBP, and DBP were 89.7±8.7 mmHg, 124.1±11.2 mm Hg, and 73.3±8.7 mm Hg, respectively. NSAID treatment affected MAP and SBP over the 12-hour dosing interval (Figure 2). Naproxen treatment significantly increased MAP (Difference in LS means: 3.1 mm Hg (95% CI: 2.1, 4.1); p<0.05), SBP (Difference in LS means: 2.9 mm Hg (95% CI: 1.8, 4.0); p<0.05) and DBP (Difference in LS means: 3.2 mm Hg (95% CI: 2.2, 4.2); p<0.05) relative to placebo. In contrast, celecoxib treatment did not affect MAP (Difference in LS means: 0.6 mm Hg (95% CI: -0.3, 1.6); p=0.28), but significantly decreased SBP (Difference in LS means: -1.1 mm Hg (95% CI: -2.2, 0.04); p<0.05) and increased DBP (Difference in LS means: 1.4 mm Hg (95% CI: 0.4, 2.4); p<0.05) relative to placebo. Compared to celecoxib, naproxen significantly increased MAP (Difference in LS means: 2.5 mm Hg (95% CI: 1.5, 3.5); p<0.05), SBP (Difference in LS means: 4.0 mm Hg (95% CI: 2.9, 5.1); p<0.05), and DBP (Difference in LS means: 1.8 mm Hg (95% CI: 0.8, 2.8); p<0.05).

**Figure 2.**
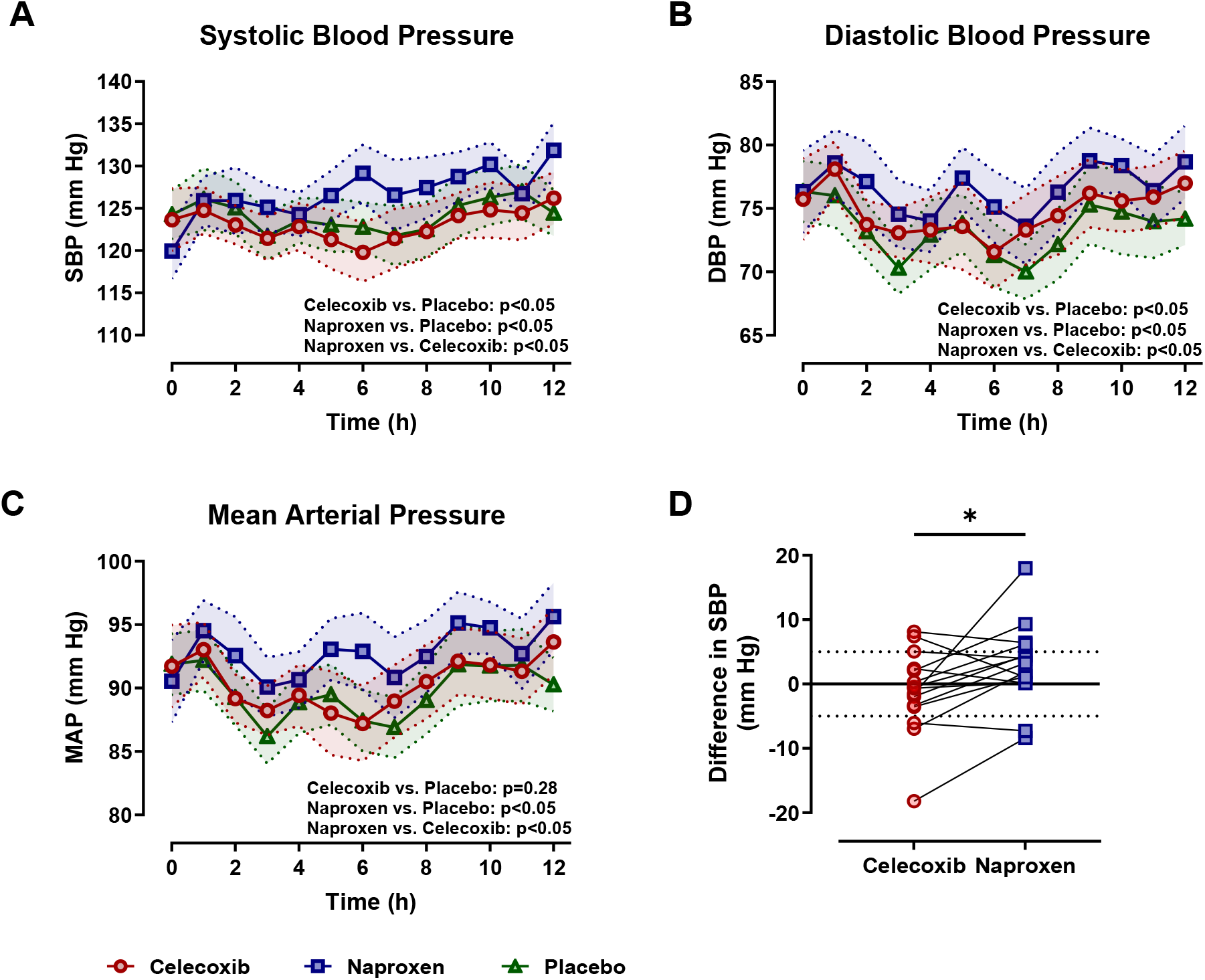
Comparison of (A) SBP, (B) DBP, and (C) MAP over 12-hour dosing interval by treatment. Data are shown as mean ± SEM. (D) Change in SBP relative to placebo with celecoxib and naproxen treatment. Dosing occurred at 8:00 AM ± 15 min. *p<0.05

Linear regression was performed to determine whether the degree of COX-1 or COX-2 inhibition *ex vivo* predicted the difference in SBP with NSAID treatment relative to placebo (Figure 3). COX-2 inhibition *ex vivo* was a predictor of difference in SBP (β=-10.38, 95% confidence interval=-19.27 to -1.482, R^2^=0.1694, F(1,28)=5.710, p=0.0238), but COX-1 inhibition *ex vivo* was not significantly associated with difference in SBP (β=-2.194, 95% confidence interval=-6.068 to 1.680, R^2^=0.04587, F(1,28)=1.346, p=0.2558).

**Figure 3.**
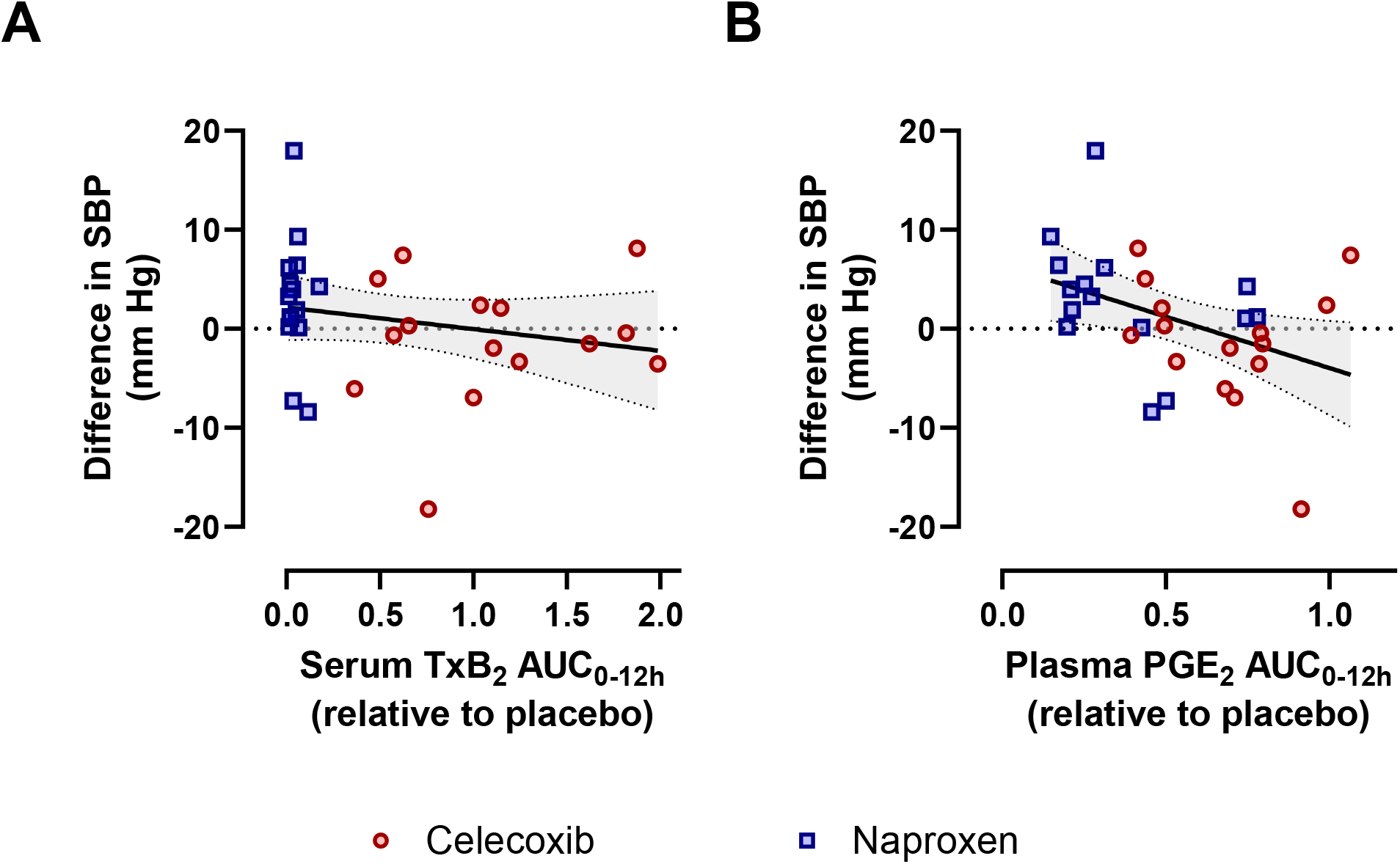
Relationship between degree of COX-1 and COX-2 inhibition *ex vivo* and change in systolic blood pressure.

## Discussion

Given the high prevalence of chronic pain in the United States,^26^ NSAIDs are an important non-addictive option for pain relief.^2^ Currently, it is recommended that NSAIDs be avoided or used only for a limited duration at the lowest possible dose in patients considered at high cardiovascular risk.^27^ As COX-2 inhibition mechanistically underlies the cardiovascular risk, a key question remains whether differences in the cardiovascular safety profile exist between traditional NSAIDs (i.e. naproxen) which inhibit both COX-1 and COX-2, and selective inhibitors of COX-2 (i.e. celecoxib).^28^ The largest comparator trial to address this question, PRECISION, indicated that celecoxib is noninferior to naproxen and ibuprofen with regard to cardiovascular risk.^17^ Importantly, the dose of celecoxib was limited to 200 mg/day in osteoarthritis patients enrolled in PRECISION, which made up the vast majority of the study population. Limiting the dose in osteoarthritis patients had been a regulatory response to the cardiovascular hazard detected in previous randomized controlled trials. Although the pharmacological potency of celecoxib measured with isolated COX-2 enzyme or cellular preparations *in vitro* is higher than that of naproxen,^29^ here we demonstrate that the average daily dose of celecoxib used in the PRECISION trial inhibited COX-2 activity *in vivo* to a lesser degree than a low dose of naproxen and that this impacted the blood pressure response to NSAID treatment. Our results underscore the importance of considering dose and pharmacoequivalence *in vivo* in comparisons of safety among drugs of the same class.

Only a small number of clinical trials have prospectively assessed the effects of COX inhibition on BP control. Based on these studies, increases of 3-5 mmHg in SBP can be expected.^16^ In our cohort, naproxen treatment increased SBP relative to placebo (2.9 mm Hg (95% CI: 1.8,4.0)) to a greater extent than celecoxib treatment (-1.1 mm Hg (95% CI: -2.2,-0.04)) over the 12-hour dosing interval. This difference is similar to what was observed in PRECISION-ABPM, where the change in SBP from baseline after 4 months of treatment was - 0.18±9.400 mm Hg among celecoxib-treated patients and 1.91±9.796 mm Hg among naproxen-treated patients.^19^ Notably, we observed that the difference in SBP was associated with the degree of COX-2 inhibition on NSAID treatment, consistent with the inhibition of COX-2-mediated PGI_2_ formation as the primary mechanism underlying the increased cardiovascular risk associated with NSAID use.

Although naproxen increased SBP to a greater extent than celecoxib in our study cohort, the blood pressure response to NSAID treatment was heterogeneous among individual patients. The effects of COX inhibition on blood pressure control are complex, which may contribute to the variable occurrence of hypertension on NSAIDs.^10,15,16,30^ In the renal cortex, the production of vasodilatory PGE_2_ and PGI_2_ maintain the patency of adjacent afferent arterioles,^16,31^ and COX-2 expression in renal medullary interstitial cells play an important role in the adaptive regulation of blood pressure in response to high salt diet and dehydration.^32-34^ COX inhibition in these regions of the kidney contributes to the decline in glomerular filtration rate and elevations in blood pressure observed in patients who take NSAIDs. However, dynamic expression of COX-2 in the macula densa system is a component of the tubuloglomerular feedback mechanism which promotes renin release.^35-37^ Inhibition of COX-2 in these cells would counteract the hypertensive effects of renin-angiotensin system activation. Thus, the effect of NSAID treatment on blood pressure reflects the complex interplay among these regulatory systems. Interestingly, we observed that an individual participant’s response was similar to both drugs, suggesting that patient-specific factors may contribute to interindividual heterogeneity in the response to NSAIDs. Future studies are necessary to elucidate the factors that contribute to an individual patient’s risk of hypertension and other cardiovascular adverse effects with NSAID treatment.

There are limitations to our study. The small sample size limits our ability to comprehensively investigate the factors that contribute to the blood pressure response to NSAIDs. The study cohort included only healthy adults, most of whom were relatively young. Although this limits potential confounding due to effects of age and comorbidities, it precludes interrogation of the influence of these factors on the blood pressure response to NSAID treatment. Finally, we compared only one dose level of naproxen and celecoxib, which limits our ability to extrapolate our results to higher doses or other NSAIDs. Despite these limitations, our results provide mechanistic insight into the outcome of PRECISION.

In conclusion, our results demonstrate that naproxen 500 mg/day inhibits COX-2 activity to a greater degree than celecoxib 200 mg/day and that the degree of COX-2 inhibition is associated with the blood pressure response to NSAID treatment. While PRECISION concluded non-inferiority of celecoxib compared to naproxen with regard to cardiovascular risk, this is based on a comparison of doses that are not equipotent.

## Data Availability

Data available on request due to privacy/ethical restrictions.

## Acknowledgements

Dr. FitzGerald is the McNeil Professor of Translational Medicine and Therapeutics and held a Merit Award from the American Heart Association. C.S. is the Robert L. McNeil Jr. Fellow in Translational Medicine and Therapeutics.

